# Protection against symptomatic disease with the delta and omicron BA.1/BA.2 variants of SARS-CoV-2 after infection and vaccination in adolescents: national observational test-negative case control study, August 2021 to March 2022, England

**DOI:** 10.1101/2022.08.19.22278987

**Authors:** Annabel A Powell, Freja Kirsebom, Julia Stowe, Mary E Ramsay, Jamie Lopez-Bernal, Nick Andrews, Shamez N Ladhani

## Abstract

**Background:** Little is known about the protection following prior infection with different SARS-CoV-2 variants, COVID-19 vaccination, and a combination of the two (hybrid immunity) in adolescents.

**Methods:** We used national SARS-CoV-2 testing and COVID-19 mRNA vaccination data in England to estimate protection following previous infection and vaccination against symptomatic PCR-confirmed delta and omicron BA.1/BA.2 variants in 11-17-year-olds using a test-negative case-control design.

**Findings:** By 31 March 2022, 63.6% of 16-17-year-olds and 48.2% of 12-15-year-olds had received ≥1 COVID-19 mRNA vaccine dose.Between 08 August 2021 and 31 March 2022, 1,161,704 SARS-CoV-2 PCR-tests were successfully linked to COVID-19 vaccination status. In unvaccinated adolescents, prior infection with wildtype, alpha or delta provided greater protection against subsequent delta infection than subsequent omicron; prior omicron infection provided had the highest protection against omicron reinfection (59.3%; 95%CI: 46.7-69.0). In infection-naïve adolescents, vaccination provided lower protection against symptomatic omicron infection than delta, peaking at 64.5% (95%CI; 63.6-65.4) 2-14 days after dose two and 62.9% (95%CI; 60.5-65.1) 2-14 weeks after dose three, with rapidly waning protection after each dose. Previously infected and vaccinated adolescents had the highest protection, irrespective of primary infecting SARS-CoV-2 strain. The highest protection against omicron was observed in vaccinated adolescents with prior omicron infection, reaching 96.4% (95%CI, 84.4-99.1) at 15-24 weeks post dose two.

**Interpretation:** All variants provide some protection against symptomatic reinfection and vaccination adds to protection. Vaccination provides low-to-moderate protection against symptomatic omicron infection, with waning protection after each dose, while hybrid immunity provides the most robust protection.

**Funding:** None

**Research in context:** *Evidence before this study:* We have previously reported COVID-19 vaccine effectiveness in previously uninfected adolescents. There are, however, limited data on the protection offered by natural infection with different SARS-CoV-2 variants, and the added value of vaccination in previously-infected adolescents. Most studies have focused on adults and show significant protection from previous infection against re-infection with pre-omicron variants, but lower protection against omicron variants, with hybrid immunity providing the most robust protection.

*Added value of this study:* Using national SARS-CoV-2 testing and COVID-19 mRNA vaccination data in England, we were able to estimate protection afforded by previous infection, vaccination, and a combination of the two using a test-negative case-control design against PCR-confirmed symptomatic COVID-19. We found that protection against symptomatic infection with the delta variant was greater than protection against symptomatic omicron infection in those previously infected with wild-type, alpha or delta variants. Similar trends were observed in previously uninfected but vaccinated individuals. Prior omicron infection along with vaccination provided the greatest protection against further omicron variant infections.

*Implications of all the available evidence:* All variants provide some protection against future SARS-CoV-2 infection, as does COVID-19 mRNA vaccination. Our findings demonstrate, for the first time in adolescents, the additional protection afforded by hybrid immunity. In the context of the UK’s recent waves of omicron infections, our findings provide important evidence of only modest short-term protection against mild disease with omicron variants following vaccination. This has important implications for the consideration of future adolescent COVID-19 vaccination and booster programmes.

## Introduction

Adolescents have a lower risk of severe or fatal COVID-19 than adults.^1^ Consequently, in England and elsewhere, from December 2020 the roll-out of COVID-19 vaccines prioritised older adults, healthcare workers and high-risk adults. When vaccinating younger-groups, early reports of rare but potentially severe myocarditis following mRNA vaccination led the UK’s Joint Committee on Vaccination and Immunisation (JCVI) to only recommend one dose of an mRNA vaccine for 16-17-year-olds from 4 August 2021 and recommend against vaccinating healthy 12-15-year-olds as the margin of benefit was too small to support universal vaccination of this age-group.^2,3^ Ministers were, however, advised to seek advice evaluating the wider national context, which was outside the JCVI’s remit. An expert group recommended universal vaccination of 12-15-year-olds to prevent educational disruption from 13 September 2021.^4^ A second vaccine dose was subsequently recommended for both age-groups to be given 8-12 weeks after the first, which is consistent with the UK recommendation for adult vaccination.^5^ By 31 March 2022, 63.6% of 16-17-year-olds and 48.2% of 12-15-year-olds had received at least one COVID-19 mRNA vaccine dose in England, 42.9% and 25.7%, respectively, had received at least two doses and 6.3% and 0.2%, respectively, had received three doses.^6^

The UK has experienced multiple waves of SARS-CoV-2 infections since March 2020, often following the emergence and rapid spread of new variants, including alpha in November 2020, delta in April 2021 and omicron in November 2021. Delta was more transmissible than alpha,^7^ but, unlike in adults,^8^ was not associated with more severe disease in children and adolescents.^9^ The ability of the omicron BA.1 variant identified in England late November 2021 and, subsequently, the BA.2 variant identified late December 2021, to evade both natural and vaccine-induced immunity was associated with the highest case numbers across all age-groups, although hospitalisation rates and fatalities remained low,^10^ likely due to prior immunity from a combination of previous infections and vaccination,^11^ along with a predilection of the omicron variant to infect the upper rather than lower airway, thus causing less severe disease.^12,13^

As in adults, we and others have shown a modest reduction in vaccine effectiveness (VE) over time against symptomatic delta disease in previously-uninfected adolescents but much lower VE against symptomatic omicron disease, although protection against severe disease with both variants has remained high.^14-16^ Recent studies have shown that adults with hybrid immunity from a combination of prior infection and vaccination had greater and longer-lasting protection against re-infection compared to previously-infected unvaccinated adults or previously-uninfected adults receiving ≥2 COVID-19 vaccine doses.^17,18^ Whether the same trends occur in adolescents, who are more likely to remain asymptomatic or develop a mild illness when exposed to SARS-CoV-2, is not known.^1^

In England, SARS-CoV-2 PCR testing was freely available from June 2020 to March 2022. Given that around half the adolescents have had ≥1 COVID-19 vaccine dose since September 2021, along with high levels of community PCR-testing, this age-group provides a unique opportunity to assess protection from previous infection, mRNA vaccination and hybrid immunity in adolescents.

## Methods

We used a test-negative case-control design to estimate protection against PCR-confirmed COVID-19 (symptomatic SARS-CoV-2 infection) after combinations of previous infection with wild type, alpha, delta and omicron variants of SARS-CoV-2 with one or two BNT162b2 (Comirnaty, Pfizer-BioNTech) doses or a booster dose of either BNT162b2 or mRNA-1273 (Spikevax, Moderna) in England during periods of delta or omicron dominance (Supplement 1). Detailed methodology used to estimate VE in previously uninfected individuals has been published.^19,20^ Vaccination status in symptomatic 12-17-year-olds with PCR-confirmed COVID-19 was compared with vaccination status in symptomatic adolescents who had a negative SARS-CoV-2 PCR test.

### Data sources

Sources of data on vaccination status, testing, identification of variants, covariates included in the analyses and data linkage methods have been described previously.^19,20^ PCR-testing data for adolescents were extracted on 31/05/2022 from 09/08/2021 when routine vaccination uptake for 16-17-year-olds started to increase to 31/03/2022, when community SARS-CoV-2 PCR-testing ended, and linked to National Immunisation Management Service (NIMS) on 31/05/2022 using combinations of unique individual National Health Service (NHS) number, date of birth, surname, first name, and postcode using deterministic linkage.

### SARS-CoV-2 testing data

Symptomatic adolescents who had a SARS-CoV-2 PCR test in the community (Pillar 2) were included in the analysis. Previous infection was defined as PCR-confirmed SARS-CoV-2 infection occurring ≥90 days before current sample date. Negative tests taken within 7 days of a previous negative test, and negative tests where the symptom onset date was within the 10 days of a symptom onset date for a previous negative test, were dropped as these likely represent the same episode. Negative tests taken within 21 days of a subsequent positive test were excluded as chances are high that these are false negatives. Positive and negative tests within 90 days of a previous positive test were excluded; however, where participants had later positive tests within 14 days of a positive test, then preference was given to PCR tests and tests associated with symptomatic infection. For adolescents who had more than one negative test, a maximum of two negative tests could be included per person, one from before 22/11/2021 and one after 22/11/2021 (before and after omicron emergence, respectively). Data were restricted to persons who reported symptoms and gave a symptom onset date within the 10 days before testing to account for reduced PCR sensitivity beyond this period. Only those unvaccinated at symptom onset and primary immunisation with BNT162b2 at symptom onset were included.

### Identification of Variants and Assignment to Cases

The variant responsible for each case was defined according to whole genome sequencing, genotyping, S-gene target failure (SGTF) status, or time period, with sequencing taking priority, followed by genotyping followed by SGTF status, as described previously.^19,20^ Where subsequent positive tests within 14 days included sequencing, genotyping or SGTF information, this information was also used to classify the variant. S target–negative status was used to define the omicron variant when it was responsible for at least 80% of S target–negative cases. From January 10, 2022, delta cases were identified by sequencing and genotyping because the positive predictive value of S target-negative status to identify the delta variant had decreased and could no longer be used. Tests were defined as delta where there was no SGTF until week 47 2021, and as omicron where there was no SGTF from week 1 2022 (Supplement 2). “Omicron” includes BA.1 and BA.2 subvariants which were both circulating widely during the study period. We did not separate BA.1/BA.2 in our analysis because the two subvariants appeared in quick succession over a short time period,^21^ (supplement 2) and our previous analysis of real-world data showed similar VE, and rate of decline of VE over time between the two subvariants in adults.^22^

Where there was a previous positive test at ≥90 days before the current test, the variant of that test was assigned based only on the dominant strain of the period in which that test occurred. For those with more than one previous positive, the first positive test was used. Wild type was assumed for the period before 8/12/2020, alpha for 08/12/2020 – 09/05/2021, delta for 10/05/2021-12/12/2021, and omicron from 12/12/2021.

### Statistical analysis

We have previously reported the methodology for test-negative case-control analysis in previously uninfected individuals.^19,20,23^ Logistic regression was performed, with the PCR-test result as the dependent variable and vaccination status as an independent variable. VE/protection was defined as 1-odds of vaccination in cases/odds of vaccination in controls, with vaccination stratified by time interval after each dose and previous variant infection. The effectiveness of combinations of vaccination and past infection was adjusted for in logistic regression models of age, sex, index of multiple deprivation (quintile), ethnic group, geographic region (NHS region), period (calendar week of test), clinical risk group status (a separate flag for those aged over and under 16 years), and clinically extremely vulnerable (if aged 16 and above). Age was defined as at 31/08/2021. Vaccination periods considered after each dose were 0-1 week, 2-14 weeks, 15-24 weeks, 25-39 weeks and 40+ weeks. For booster vaccination, data for BNT162b2 or mRNA-1273 vaccinations were combined. Infection status was determined relative to 7 days after the first vaccination dose, with those infected before this regarded as infected prior to vaccination, and those infected after regarded as infected after vaccination (irrespective of timing of other doses).

## Results

Between 09 August 2021 and 31 March 2022, there were 1,161,704 tests successfully linked to NIMS for COVID-19 vaccination status, including 390,467 positive tests against the delta variant and 212,433 positive tests against the omicron variant (Supplement 3,4&5). Overall, there were 558,804 negative tests, which included 460,756 tests during the delta period and 225,447 during omicron. Some negative tests could be used as controls for both delta and omicron periods. Characteristics and vaccination status of adolescents tested are summarised in Supplement 3.

### Protection from past infection in unvaccinated adolescents

In unvaccinated adolescents, protection against symptomatic delta infection was 87.6% (95%CI; 86.8-88.4) after previous confirmed wildtype infection, 86.1% (95%CI; 85.4-86.8) after alpha and 92.3% (95%CI; 91.7-92.9) after delta (**Table 1**). For symptomatic omicron infection, protection was 32.7% (95%CI; 27.7-37.4), 36.6% (95%CI; 32.9-40.1) and 52.4% (95%CI; 50.9-53.8) respectively (**Table 1**). Prior omicron infection was 59.3% (95%CI; 46.7-69.0) protective against omicron re-infection, which is significantly lower than delta infection against delta re-infection (92.3%, 95%CI; 91.7-92.9).

**Table 1.** Effectiveness of past infection by variant against symptomatic infection with the delta and omicron variants in unvaccinated adolescents.

| Past infection | Delta<br>*VE (95%CI)* | Omicron<br>*VE (95%CI)* |
| --- | --- | --- |
| Wild type | 87.6 (86.8-88.4) | 32.7 (27.7-37.4) |
| Alpha | 86.1 (85.4-86.8) | 36.6 (32.9-40.1) |
| Delta | 92.3 (91.7-92.9) | 52.4 (50.9-53.8) |
| Omicron | - | 59.3 (46.7-69.0) |

### Protection against delta

Overall, mRNA vaccination protected against symptomatic delta infection in infection-naïve adolescents and provided additional protection in previously infected adolescents. In infection-naïve adolescents, VE against delta after one vaccine dose was 59.4% (95%CI; 58.8-60.0) after 2-14 weeks but dropped to 23.5% (95%CI; 18.3-28.3) after 15-24 weeks (**Table 2, Figure 1**). After two doses, VE increased to 91.8% (95%CI; 91.2-92.3) after 2-14 weeks before falling to 71.9% (95%CI; 67.9-75.4) after 25-29 weeks. A third dose increased VE to 96.0% (95%CI; 92.2-97.9) after 2-14 weeks. Further follow-up was limited by the emergence of the omicron variant.

**Table 2.** Protection of the combinations of vaccination and previous infection with wild type, alpha, delta and omicron variants of SARS-CoV-2 against the delta variant by weeks since vaccination.

|  |  | Previous infection (before vacc) |  |  |  | Previous infection (after vacc) |
| --- | --- | --- | --- | --- | --- | --- |
|  |  | None | Wild type | Alpha | Delta | Delta |
| Vaccination status | Time after vaccination | VE (95% CI) | VE (95% CI) | VE (95% CI) | VE (95% CI) | VE (95% CI) |
| Unvaccinated |  | Baseline | 87.6 (86.8-88.4) | 86.1 (85.4-86.8) | 92.3 (91.7-92.9) | 92.3 (91.7-92.9) |
| Dose 1 - Pfizer | 0-1 week | 8.5 (6.7-10.3) | 92.6 (90.4-94.3) | 90.3 (88.4-92.0) | 91.3 (89.2-93.0) |  |
|  | 2-14 weeks | 59.4 (58.8-60.0) | 98.1 (97.6-98.6) | 95.5 (94.8-96.1) | 97.5 (97.0-97.9) | 95.2 (64.5-99.4) |
|  | 15-24 weeks | 23.5 (18.3-28.3) | 98.6 (90.3-99.8) | 94.2 (85.9-97.6) | 99.0 (92.8-99.9) | 98.4 (88.3-99.8) |
|  | 25-39 weeks | 57.4 (39.0-70.2) | - | - | - | - |
|  | 40 weeks |  | - | - | - | - |
| Dose 2 - Pfizer | 0-1 week | 71.0 (68.9-73.0) | - | 98.4 (95.0-99.5) | 99.6 (97.1-99.9) | 94.2 (76.3-98.6) |
|  | 2-14 weeks | 91.8 (91.2-92.3) | 98.8 (96.7-99.5) | 99.2 (97.8-99.7) | 98.7 (96.8-99.4) | - |
|  | 15-24 weeks | 80.9 (79.4-82.3) | 98.6 (94.3-99.7) | 97.0 (92.7-98.8) | - | 98.3 (87.9-99.8) |
|  | 25-39 weeks | 71.9 (67.9-75.4) | 94.8 (78.4-98.8) | 97.3 (80.2-99.6) | - | 96.4 (73.2-99.5) |
|  | 40 weeks |  | - | - | - | - |
| Booster - Any mRNA vaccine | 0-1 week | 84.8 (77.6-89.7) | - | - | - | - |
|  | 2-14 weeks | 96.0 (92.2-97.9) | - | - | - | - |
|  | 15-24 weeks |  | - | - | - | - |

**Figure 1.**
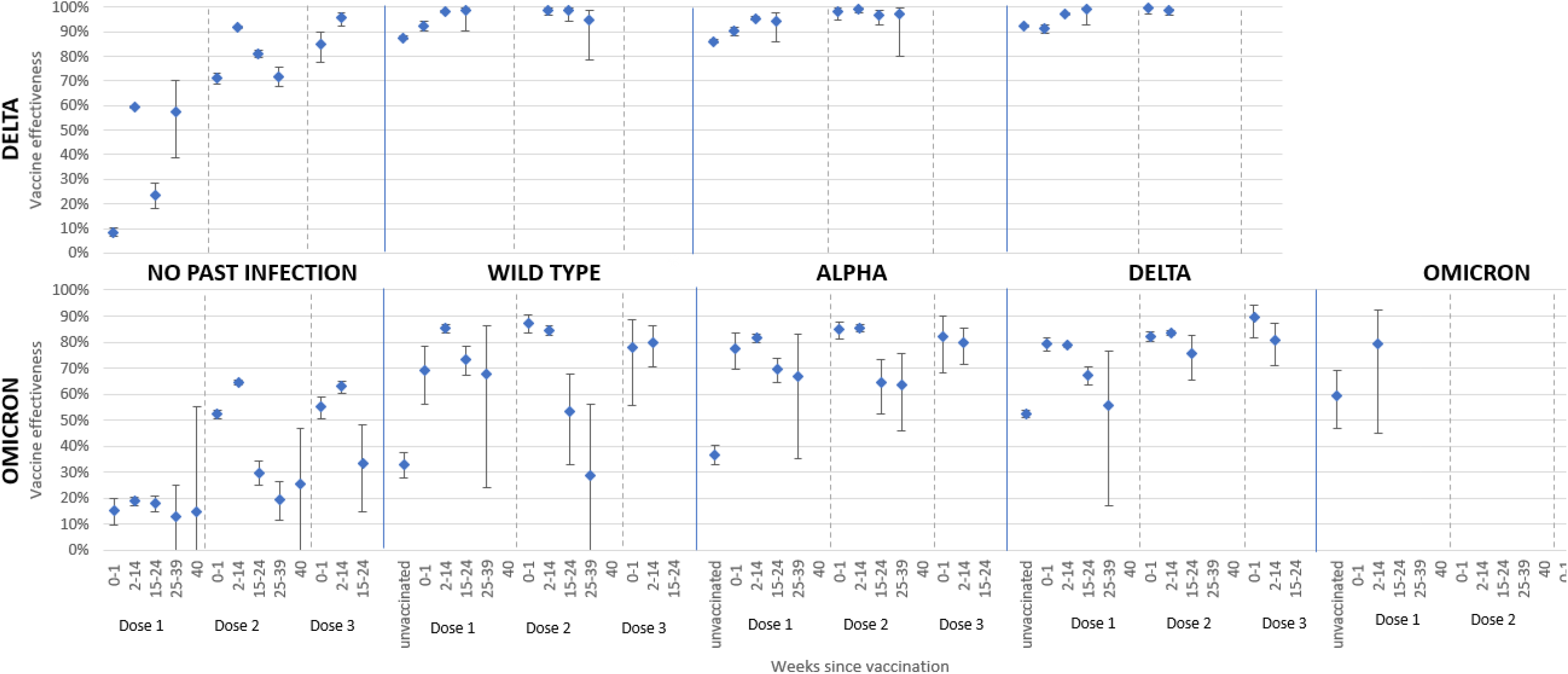
Protection of the combinations of vaccination and previous infection with wild type, alpha, delta and omicron variants of SARS-CoV 2 by weeks since vaccination.

In previously infected adolescents, protection against symptomatic delta infection was above 90% with limited waning over time after vaccination, irrespective of the infecting strain (wildtype, alpha or delta). Protection against symptomatic delta re-infection remained high and above 90% among adolescents irrespective of whether the infection occurred before or after vaccination (**Table 2, Figure 2**).

**Figure 2.**
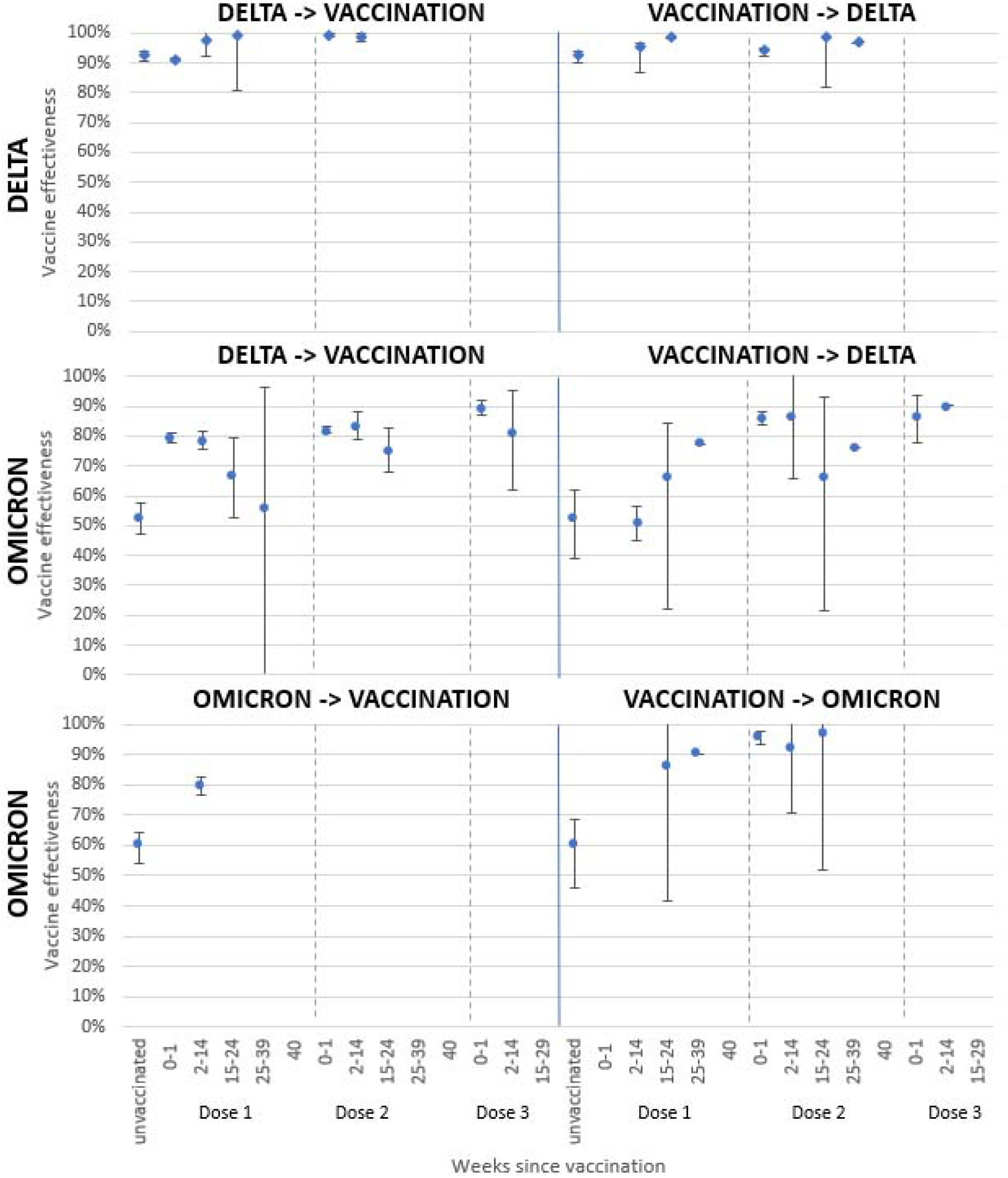
Protection of the combinations of vaccination according to SARS-CoV-2 infection then vaccination or vaccination then SARS-CoV-2 infection with delta or omicron variants against delta or omicron re-infection by weeks since vaccination.

### Protection against omicron

Overall, mRNA vaccination protected against symptomatic omicron infection in infection-naïve adolescents and provided additional protection in previously infected adolescents, but to a lower extent than against delta (**Figure 1, Table 3**). In infection-naïve adolescents, VE against omicron was 18.8% (95%CI; 17.2-20.3) 2-14 weeks after dose one and peaked at 64.5% (95%CI; 63.6-65.4) 2-14 weeks after dose two before declining to 19.4% (95%CI; 11.7-26.4) by 25-39 weeks (**Figure 1, Table 3**). VE after three doses increased to 62.9% (95%CI; 60.5-65.1) after 2-14 weeks and then declined.

**Table 3.** Protection of the combinations of vaccination and previous infection with wild type, alpha, delta and omicron variants of SARS-CoV-2 against the omicron variant by weeks since vaccination.

| Vaccination status | Time after vaccination | Previous infection (before vacc) |  |  |  |  | Previous infection (after vacc) |  |
| --- | --- | --- | --- | --- | --- | --- | --- | --- |
|  |  | None | Wild type | Alpha | Delta | Omicron | Delta | Omicron |
|  |  | VE (95% CI) | VE (95% CI) | VE (95% CI) | VE (95% CI) | VE (95% CI) | VE (95% CI) | VE (95% CI) |
| Unvaccinated |  | baseline | 32.7 (27.7-37.4) | 36.6 (32.9-40.1) | 52.4 (50.9-53.8) | 59.3 (46.7-69.0) | 52.4 (50.9-53.8) | 59.3 (46.7-69.0) |
| Dose 1 - Pfizer | 0-1 week | 15.2 (9.9-20.1) | 69.2 (55.9-78.5) | 77.6 (69.5-83.6) | 79.3 (76.7-81.6) |  |  |  |
|  | 2-14 weeks | 18.8 (17.2-20.3) | 85.3 (83.7-86.8) | 81.5 (80.0-82.9) | 78.8 (77.9-79.5) | 79.6 (44.9-92.4) | 51.2 (28.4-66.8) |  |
|  | 15-24 weeks | 17.9 (14.9-20.7) | 73.4 (67.2-78.4) | 69.5 (64.5-73.8) | 67.2 (63.7-70.3) |  | 65.9 (60.5-70.6) | 85.5 (77.5-90.6) |
|  | 25-39 weeks | 12.8 (-1.6-25.1) | 67.8 (24.1-86.3) | 66.7 (35.2-82.9) | 55.8 (17.2-76.4) |  | 77.2 (67.5-84) | 90.2 (75.9-96.0) |
|  | 40 weeks | 14.7 (-63.3-55.4) |  |  |  |  |  |  |
| Dose 2 - Pfizer | 0-1 week | 52.2 (50.4-53.9) | 87.4 (83.5-90.4) | 84.9 (81.3-87.8) | 82.1 (80.1-83.9) |  | 86.1 (82.3-89.1) | 95.5 (85.3-98.6) |
|  | 2-14 weeks | 64.5 (63.6-65.4) | 84.7 (82.6-86.5) | 85.5 (84.0-86.9) | 83.5 (82.5-84.5) |  | 86.5 (85.1-87.8) | 91.2 (80.0-96.1) |
|  | 15-24 weeks | 29.8 (24.9-34.2) | 53.4 (32.7-67.7) | 64.3 (52.4-73.3) | 75.5 (65.6-82.5) |  | 65.9 (55.2-74.1) | 96.4 (84.4-99.1) |
|  | 25-39 weeks | 19.4 (11.7-26.4) | 28.9 (-15.5-56.3) | 63.6 (46.0-75.5) |  |  | 76.1 (65.3-83.6) |  |
|  | 40 weeks | 25.7 (-4.2-47.0) |  |  |  |  |  |  |
| Booster - Any mRNA vaccine | 0-1 week | 55.1 (50.7-59.1) | 77.7 (55.7-88.8) | 82.2 (68.1-90.1) | 89.5 (81.7-94.0) |  | 87 (78.8-92.0) |  |
|  | 2-14 weeks | 62.9 (60.5-65.1) | 79.8 (70.4-86.3) | 79.6 (71.4-85.5) | 80.7 (71.1-87.1) |  | 90.1 (86.6-92.7) |  |
|  | 15-24 weeks | 33.6 (14.6-48.3) |  |  |  |  |  |  |

Vaccination against symptomatic omicron infection in adolescents previously infected with wildtype, alpha, delta or omicron variants increased protection, with higher peaks compared to infection-naïve adolescents, reaching 85.3% (95%CI; 83.7-86.8), 81.5% (95%CI; 80.0-82.9), 78.8% (95%CI; 77.9-79.5) and 79.6% (95%CI; 44.9-92.4), respectively, 2-14 weeks after one dose (**Figure 1, Table 3**).

Protection waned after the first dose and then increased to 80-90% 2-14 weeks after the second dose before waning again, being more evident in adolescents previously infected with wildtype, reaching a nadir of 28.9% (95%CI; -15.5-56.3), albeit with wide confidence intervals, compared to those previously infected with alpha (63.6%, 95%CI; 46.0-75.5) or delta (75.5%, 95%CI; 65.6-82.5) at 25-29 weeks after dose two. (**Figure 1, Table 3**) A third vaccine dose boosted protection to 80-90% in adolescents previously infected with wildtype, alpha or delta variant, but follow-up was limited to 2-14 weeks after dose three.

Similar trends against symptomatic omicron infection were observed for adolescents who were infected with delta before vaccination compared to those with delta infection after vaccination (**Figure 2, Table 3**).

There was limited follow-up for adolescents vaccinated after omicron infection, with wide confidence intervals because of small case numbers (**Figure 1, Table 3**). Adolescents who were vaccinated before omicron infection showed the highest protection against symptomatic omicron re-infection with protection remaining at 90.2% (95%CI; 75.9-96.0) 25-39 weeks after dose one and 96.4% (95%CI; 84.4-99.1) at 15-24 weeks after dose two (**Figure 2, Table 3**).

## Discussion

There are limited data on the effects of natural and vaccine-induced immunity against SARS-CoV-2 in adolescents, with most reported studies focussing on adults. We found that primary SARS-CoV-2 infection with wildtype (88%), alpha (86%) or the delta (92%) variant was highly protective against subsequent symptomatic delta infection in unvaccinated adolescents, but less so against symptomatic omicron infection (33%, 37% and 52% respectively), while prior omicron infection provided 59% protection against omicron re-infection. In infection-naïve adolescents, two mRNA vaccine doses provided 92% protection against delta, although protection waned with time, high VE was restored after a third dose. In contrast, two doses provided lower protection (65%) against symptomatic omicron infection, with rapidly waning protection after each dose and a similar trend observed after a third dose. In previously infected adolescents, vaccination with one or two doses provided high protection against delta irrespective of the SARS-CoV-2 variant responsible for primary infection, which was sustained after each vaccine dose, with very little waning. Vaccination also added protection against symptomatic omicron infection in previously infected adolescents, but with a lower peak and greater waning after each vaccine dose. High protection was observed against symptomatic omicron re-infection in vaccinated adolescents who were previously infected with omicron.

### Prior infection

Our findings confirm studies in adults showing significant protection from previous infection against re-infection with pre-omicron variants.^17,24^ A recent Qatari study reported 90% protection of previous infection against delta re-infection and 62% against omicron in unvaccinated adults.^24^ This is consistent with our data with unvaccinated adolescents showing lower protection against reinfection with omicron compared to delta. This is likely as omicron variants harbour critical mutations leading to evasion of natural and vaccine-induced immunity.^25^ Reassuringly, adult studies show similar protection by mRNA vaccines against both BA.1 and BA.2 subvariants.^22^

### Vaccination alone

As in adults,^14,26^ we found limited, short-term protection from vaccination against symptomatic omicron infection, especially when compared to delta in infection-naïve adolescents. We have previously reported higher VE in 12-15- and 16-17-year-olds against symptomatic delta compared to omicron, including rapid waning after each dose against symptomatic omicron infection in 16-17-year-olds which is consistent with our recent analysis.^14^ Here, we show that VE against symptomatic omicron infection wanes rapidly after each vaccine dose in infection-naïve adolescents and remains low at 33% even after three vaccine doses. In a recent Qatari adult study, there was no protection against omicron BA.2 from six months after two BNT162b2 doses and 52% protection ∼43 days after three doses.^18^ Although timepoints are not directly comparable, consistent with Qatari data, we also see a significant drop in VE to 19% 25-39 weeks after dose two and 63% 2-14 weeks after dose three.^18^

### Hybrid immunity

Consistent with emerging literature in adults,^17,18,24^ and laboratory data in children and adolescents,^27^ we found that hybrid immunity provided the most robust protection. In our cohort, protection against the delta variant remained above 90% after prior infection with any pre-omicron variant and one or two BNT162b2 doses. Whilst vaccination improved protection against omicron in previously infected adolescents, peak protection remained lower than against delta, with substantial waning over time after the first and second vaccine dose, although protection remained at 80-90% up to three months after the third dose. These data are consistent with the Qatari study where, against BA.2 infection, previous infection and two BNT162b2 doses was 55.1% (95%CI, 50.9-58.9) protective at a median of 270 days between the second dose and PCR test, while previous infection and three BNT162b2 doses being 77.3% (95%CI, 72.4-81.4) protective at a median of 43 days between the third dose and PCR test.^18^ These findings are consistent with laboratory studies showing that hybrid immunity provides broader protection with higher neutralising antibody activity and greater protection against SARS-CoV-2 compared to infection or vaccination alone.^28^

### Waning

Waning of SARS-CoV-2 antibodies has been well-documented after natural infection and vaccination. Indeed antibody waning is considered the main reason for recurrent infections with seasonal coronaviruses.^29,30^ We also observed rapid waning of immunity against infection offered by vaccination in infection-naïve persons, consistent with adult studies.^17,31,32^ Among previously infected individuals, we found large disparities with delta and omicron, with high protection against symptomatic delta infection maintained even after one vaccine dose. In contrast, vaccination in previously infected adolescents achieved lower peak protection against symptomatic omicron infection and greater waning of protection after each vaccine dose compared to delta. Notably, though, vaccination then omicron infection provided the most robust protection against omicron reinfection, although we were unable to assess duration of protection because of limited follow-up time. As with delta, it is likely that omicron infection followed by vaccination would also provide robust protection, but we did not have sufficient data to assess this in our cohort. The sequence of infection and vaccination is likely less important, with protection against re-infection following primary infection then vaccination being similar to vaccination then primary infection, especially against delta but also against omicron.^17^

### Severe disease

Our analysis did not assess protection against severe disease, hospitalisations or deaths, which has been demonstrated for adults, these show despite waning immunity against symptomatic infection over time and with the emergence of new variants, protection against severe disease remains high and long-lasting.^24,33^

We and others have demonstrated vaccination offering significant protection against hospitalisation in previously-uninfected adolescents.^14-16^ Given the high prevalence of asymptomatic infection,^34^ and the low risk of severe COVID-19,^1^ along with milder disease associated with omicron,^10^ it will become more difficult to compare differences in protection from prior infection with different variants alongside different vaccine brands and number of vaccine doses. Using hospitalisations as a marker of severe COVID-19 will also be difficult because of disproportionately higher hospitalisation rates among those with underlying comorbidities compared to healthy adolescents, and high rates of incidental infections among the hospitalised during periods of high community infection rates.^35^

### Implications

Our population-based analysis of protection against symptomatic infection after natural, vaccine-induced and hybrid immunity in adolescents is consistent with adult studies. With the high case rates observed with omicron globally and emergence of multiple immune-evasive omicron subvariants, it is likely that, as in England,^36^ nearly all children have been exposed to and developed some immunity to SARS-CoV-2. Additionally, adolescents in many countries have been vaccinated against COVID-19, providing them with additional protection.

Since community testing ended in England, new subvariants of omicron, BA.4 and BA.5, have emerged, and, appear to be more evasive than previous omicron sub-variants, resulting in high rates of re-infections irrespective of previous infection or vaccination status, although hospitalisations and deaths in adults remains low.^37^ Taken together, these data question the need for COVID-19 vaccine boosters in highly immune populations.

Free community PCR or LFD tests ended in the UK in March 2022 and, therefore, analyses such as this one will no longer be possible and will be restricted to monitoring severe outcomes such as hospitalisation and death.

### Strengths and Limitations

The large scale of testing and sequencing in the UK, and the use of the national vaccination register, has allowed broad evaluation of protection in adolescents against delta and omicron, as well as evaluation of protection from previous infection and hybrid immunity. Our analysis, however, has some limitations. When assessing previous infection, the last infection episode was used; it is, therefore, possible that some individuals may have had earlier first infections that are not considered here, although numbers will likely be small because testing was so widely available and are unlikely to affect the overall results. Booster doses were given to adolescents during the omicron wave which prevented longer-term follow-up of protection against the delta variant. Since most adolescents were vaccinated before omicron began circulating, we were unable to assess protection beyond 2-14 weeks after one dose for this cohort. Some misclassifications may have occurred owing to imperfect sensitivity and specificity of PCR testing, and the use of S target–negative status to identify variants. It is possible, due to using real-world data, that differences not adjusted for in our multivariable analysis may be present between groups compared, as well as behaviours in those who were tested versus those who chose not to test when unwell. However, we expect these potential differences to be small and similar across the groups compared in this analysis.

## Conclusion

Prior infection with any SARS-CoV-2 variant provide some protection against reinfection, even against omicron, but more so against delta. mRNA COVID-19 vaccination always adds to protection, irrespective of past infection. Vaccination and previous infection provide very high protection against delta, with limited waning over time. Vaccination alone provides low-to-moderate protection against omicron, with waning protection after each dose, while hybrid immunity provides the most robust protection against both omicron and delta. Following high infection rates due to omicron subvariants, our findings question the need for additional vaccine doses in adolescents – we have shown that additional vaccine doses provide low-to-moderate added protection for a limited period against omicron. Further studies are needed to assess the risk of post-COVID symptoms after recurrent infection and new variants in vaccinated and unvaccinated children and adolescents.

## Data Availability

Applications for relevant anonymised data should be submitted to the UK Health Security Agency Office for Data Release: https://www.gov.uk/government/publications/accessing-ukhsa-protected-data/accessing-ukhsa-protected-data.

## Funding

None

## Declaration of interest

None

## Ethics approval

UKHSA has legal permission, provided by Regulation 3 of The Health Service (Control of Patient Information) Regulations 2002, to process patient confidential information for national surveillance of communicable diseases and as such, individual patient consent is not required to access records.

## Notes

### Competing Interest Statement

The authors have declared no competing interest.

### Funding Statement

This study did not receive any funding

### Author Declarations

Surveillance of coronavirus disease 2019 (Covid-19) testing and vaccination is undertaken under Regulation 3 of the Health Service (Control of Patient Information) Regulations 2002 to collect confidential patient information (www.legislation.gov.uk/uksi/2002/1438/regulation/3/made) under Sections 3(i) (a) to (c), 3(i)(d) (i) and (ii), and 3. The study protocol was subject to an internal review by the UK Health Security Agency Research Ethics and Governance Group and was found to be fully compliant with all regulatory requirements. Given that no regulatory issues were identified and that ethics review is not a requirement for this type of work, it was decided that a full ethics review would not be necessary.

