## Supplementary Material for "Protection against symptomatic disease with the delta and omicron BA.1/BA.2 variants of SARS-CoV-2 after infection and vaccination in adolescents: national observational test-negative case control study, August 2021 to March 2022, England"

**Supplement 1. Methods flow chart**

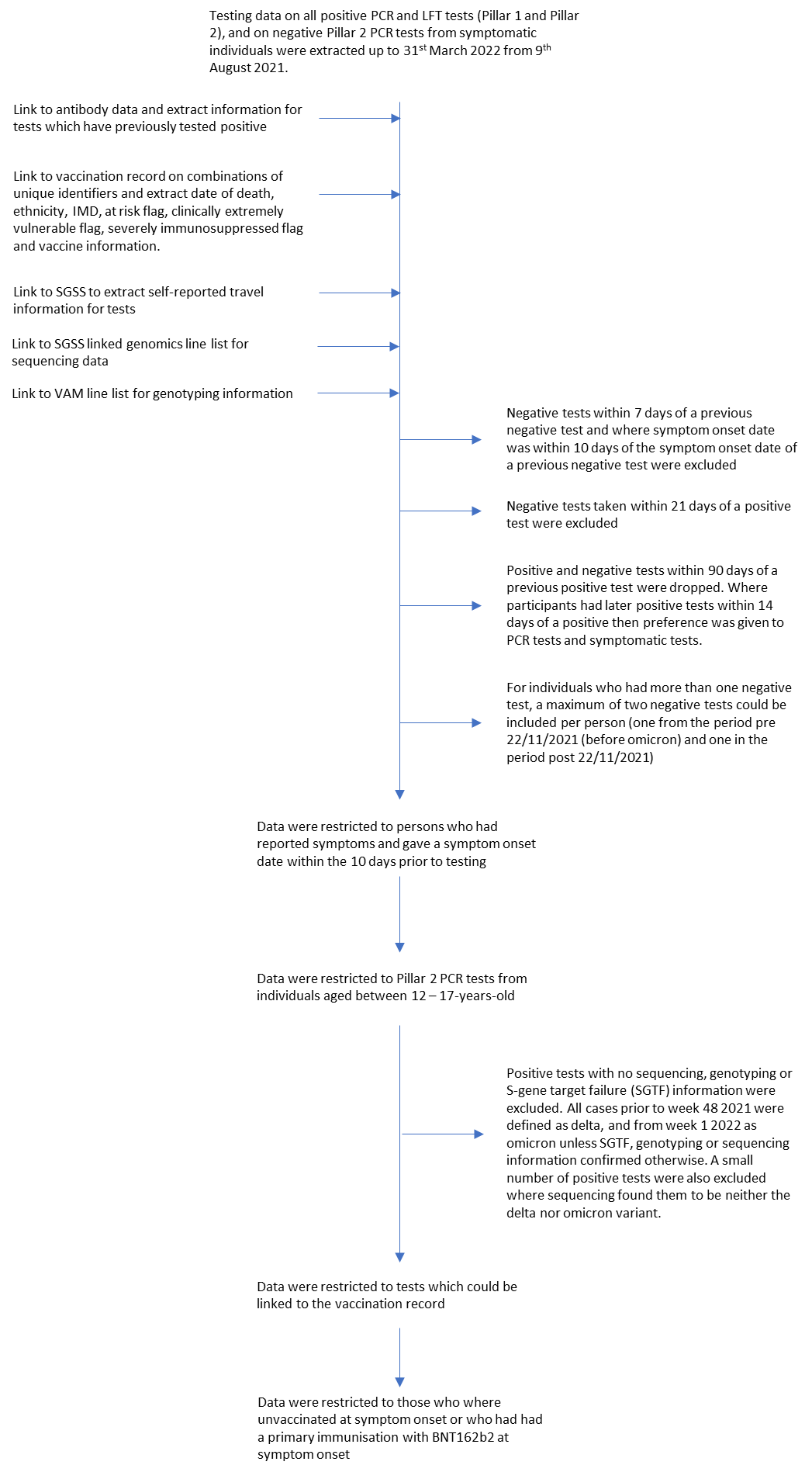

**Supplement 2. SARS-CoV-2 variant prevalence of available sequenced episodes for England from 1 February 2021 as of 18 July 2022**

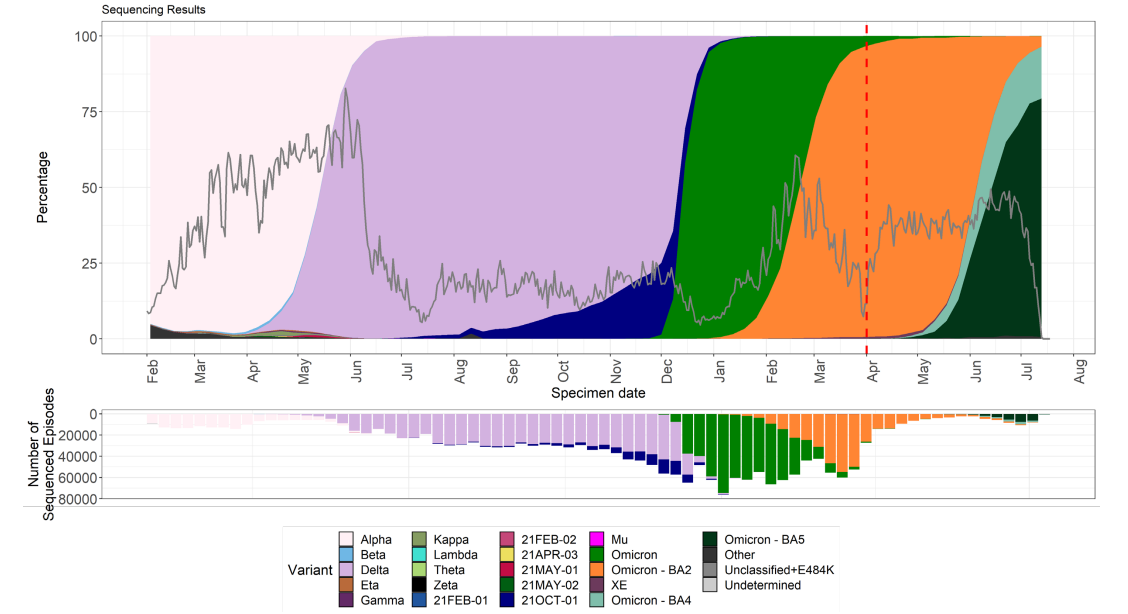

Graph taken from SARS-CoV-2 variants of concern and variants under investigation in England. Technical Briefing 44.^1^

Dashed lines indicate period incorporating issue at a sequencing site. The grey line indications proportion of cases sequenced. The red dash line denotes the start of England’s ‘Living with COVID’ plan. Note recombinants, such as XD, are not specified but are largely within the ‘other’ group currently as the numbers are too small.

Supplement 3. Descriptive characteristics of positive and negative PCR-confirmed SARS-CoV-2 test results amongst adolescents aged 12-17-years after one and two doses of BNT162b2, or three doses (BNT162b2 or m1763). Data is shown for delta and omicron.

|  |  |  | **Overall** | | **Negative** | | **Delta** | | **Omicron** | |
| --- | --- | --- | --- | --- | --- | --- | --- | --- | --- | --- |
|  |  |  | **n** | **%** | **n** | **%** | **n** | **%** | **n** | **%** |
|  | **Test Result** | **Interval (days)** | **1,161,704** |  | **558,804** |  | **390,467** |  | **212,433** |  |
| **Vaccination Status and intervals after vaccine** | Unvaccinated |  | 703,582 | 60.6 | 307,538 | 55.0 | 309,246 | 79.2 | 86,798 | 40.9 |
|  | Dose 1 - Pfizer | 0-1 week | 59,908 | 5.2 | 25,421 | 4.5 | 30,793 | 7.9 | 3694 | 1.7 |
|  |  | 2-14 weeks | 258,621 | 22.3 | 146,537 | 26.2 | 45,557 | 11.7 | 66527 | 31.3 |
|  |  | 15-24 weeks | 24,043 | 2.1 | 10,197 | 1.8 | 1,513 | 0.4 | 12333 | 5.8 |
|  |  | 25-39 weeks | 1,239 | 0.1 | 515 | 0.1 | 46 | 0.0 | 678 | 0.3 |
|  |  | 40 weeks | 60 | 0.0 | 25 | 0.0 | 1 | 0.0 | 34 | 0.0 |
|  | Dose 2 - Pfizer | 0-1 week | 21,022 | 1.8 | 12272 | 2.2 | 1112 | 0.3 | 7638 | 3.6 |
|  |  | 2-14 weeks | 70,504 | 6.1 | 43,548 | 7.8 | 948 | 0.2 | 26,008 | 12.2 |
|  |  | 15-24 weeks | 10,121 | 0.9 | 6,127 | 1.1 | 906 | 0.2 | 3,088 | 1.5 |
|  |  | 25-39 weeks | 3,891 | 0.3 | 1,937 | 0.3 | 305 | 0.1 | 1,649 | 0.8 |
|  |  | 40 weeks | 185 | 0.0 | 72 | 0.0 | 1 | 0.0 | 112 | 0.1 |
|  | Booster - Any vaccine | 0-1 week | 2,407 | 0.2 | 1263 | 0.2 | 30 | 0.0 | 1114 | 0.5 |
|  |  | 2-14 weeks | 5,809 | 0.5 | 3,229 | 0.6 | 9 | 0.0 | 2,571 | 1.2 |
|  |  | 15-24 weeks | 308 | 0.0 | 122 | 0.0 | - |  | 186 | 0.1 |
| **Age (years)** | 12 |  | 210,409 | 18.1 | 98,447 | 17.6 | 74,813 | 19.2 | 37,149 | 17.5 |
|  | 13 |  | 214,264 | 18.4 | 101,187 | 18.1 | 76,533 | 19.6 | 36,544 | 17.2 |
|  | 14 |  | 212,474 | 18.3 | 100,718 | 18.0 | 75,341 | 19.3 | 36,415 | 17.1 |
|  | 15 |  | 210,943 | 18.2 | 97,263 | 17.4 | 78,017 | 20.0 | 35,663 | 16.8 |
|  | 16 |  | 151,286 | 13.0 | 76,181 | 13.6 | 43,941 | 11.3 | 31,164 | 14.7 |
|  | 17 |  | 162,328 | 14.0 | 85,008 | 15.2 | 41,822 | 10.7 | 35,498 | 16.7 |
| **Gender** | Female |  | 597,530 | 51.4 | 282,218 | 50.5 | 200,042 | 51.2 | 115,270 | 54.3 |
|  | Male |  | 562,279 | 48.4 | 275,608 | 49.3 | 189,875 | 48.6 | 96,796 | 45.6 |
|  | Missing |  | 1,895 | 0.2 | 978 | 0.2 | 550 | 0.1 | 367 | 0.2 |
| **Ethnicity** | Asian |  | 139,768 | 12.0 | 68,172 | 12.2 | 40,735 | 10.4 | 30,861 | 14.5 |
|  | Black |  | 26,865 | 2.3 | 10,963 | 2.0 | 7,853 | 2.0 | 8,049 | 3.8 |
|  | Mixed |  | 23,670 | 2.0 | 11098 | 2.0 | 7507 | 1.9 | 5065 | 2.4 |
|  | White |  | 927,923 | 79.9 | 447,602 | 80.1 | 321,873 | 82.4 | 158448 | 74.6 |
|  | Other |  | 5,646 | 0.5 | 2,644 | 0.5 | 1,637 | 0.4 | 1365 | 0.6 |
|  | Missing |  | 37,832 | 3.3 | 18,325 | 3.3 | 10,862 | 2.8 | 8645 | 4.1 |
| **NHS Region** | East of England |  | 145,303 | 12.5 | 70,630 | 12.6 | 49,130 | 12.6 | 25,543 | 12.0 |
|  | London |  | 123,507 | 10.6 | 59,183 | 10.6 | 34,369 | 8.8 | 29,955 | 14.1 |
|  | Midlands |  | 229,938 | 19.8 | 110,088 | 19.7 | 80,117 | 20.5 | 39,733 | 18.7 |
|  | North East |  | 180,946 | 15.6 | 86,604 | 15.5 | 61,554 | 15.8 | 32,788 | 15.4 |
|  | North West |  | 152,634 | 13.1 | 75,237 | 13.5 | 47,774 | 12.2 | 29,623 | 13.9 |
|  | South East |  | 200,411 | 17.3 | 95,014 | 17.0 | 69,872 | 17.9 | 35,525 | 16.7 |
|  | South West |  | 128,958 | 11.1 | 62,045 | 11.1 | 47,648 | 12.2 | 19,265 | 9.1 |
|  | Missing |  | 7 | 0.0 | 3 | 0.0 | 3 | 0.0 | 1 | 0.0 |
| **IMD Quintiles** | 1 |  | 224,588 | 19.3 | 108,095 | 19.3 | 68,298 | 17.5 | 48,195 | 22.7 |
|  | 2 |  | 208,491 | 17.9 | 98,776 | 17.7 | 67,945 | 17.4 | 41,770 | 19.7 |
|  | 3 |  | 222,041 | 19.1 | 105,869 | 18.9 | 76,538 | 19.6 | 39,634 | 18.7 |
|  | 4 |  | 236,945 | 20.4 | 114,135 | 20.4 | 82,895 | 21.2 | 39,915 | 18.8 |
|  | 5 |  | 267,097 | 23.0 | 130,688 | 23.4 | 93,971 | 24.1 | 42,438 | 20.0 |
|  | Missing |  | 2,542 | 0.2 | 1,241 | 0.2 | 820 | 0.2 | 481 | 0.2 |
| **Vaccine priority groups** | At risk |  | 80,056 | 6.9 | 44,684 | 8.0 | 20,857 | 5.3 | 14,515 | 6.8 |
|  | CEV |  | 1802 | 0.2 | 1132 | 0.2 | 264 | 0.1 | 406 | 0.2 |
| **Previously positive** | No |  | 1,046,816 | 90.1 | 477,150 | 85.4 | 385,967 | 98.8 | 183,699 | 86.5 |
|  | Yes |  | 114,888 | 9.9 | 81,654 | 14.6 | 4,500 | 1.2 | 28,734 | 13.5 |

Negative tests are shown as totals used in analysis. As there was a period of overlap between delta and omicron, some negative tests were used in analysis for both variants.

Supplement 4. Number of positive and negative PCR-confirmed SARS-CoV-2 test results in individuals tested for SARS-CoV-2 for 12-17-year-olds in England by vaccination status, interval after vaccination and previous infection status. Data is shown for the delta variant.

|  |  | Previous infection (before vacc) | | | |  | Previous infection (after vacc) |
| --- | --- | --- | --- | --- | --- | --- | --- |
|  |  | None | Wild type | Alpha | Delta |  | Delta |
| Vaccination status | Time after vaccination | case:control | case:control | case:control | case:control |  | case:control |
| Unvaccinated |  | 251730:305486 | 7522:1092 | 12000:1951 | 9567:717 |  | 9567:717 |
| Dose 1 - Pfizer | 0-1 week | 21405:30497 | 626:67 | 938:135 | 1003:94 |  | - |
|  | 2-14 weeks | 104515:45155 | 3067:58 | 4677:200 | 7446:143 |  | 31:1 |
|  | 15-24 weeks | 3987:1505 | 155:1 | 231:5 | 281:1 |  | 131:1 |
|  | 25-39 weeks | 133:45 | 3:0 | 10:1 | - |  | 8:0 |
|  | 40 weeks | 9:1 | - | - | - |  | 1:0 |
| Dose 2 - Pfizer | 0-1 week | 5380:1106 | 185:0 | 276:3 | 506:1 |  | 109:2 |
|  | 2-14 weeks | 14626:935 | 497:4 | 762:4 | 957:5 |  | 362:0 |
|  | 15-24 weeks | 4407:897 | 134:2 | 170:5 | 14:0 |  | 78:1 |
|  | 25-39 weeks | 1348:301 | 48:2 | 55:1 |  |  | 57:1 |
|  | 40 weeks | 15:1 | - | - | - |  | 1:0 |
| Booster - Any mRNA vaccine | 0-1 week | 447:29 | 12:1 | 17:0 | 1:0 |  | 29:0 |
|  | 2-14 weeks | 674:9 | 23:0 | 21:0 |  |  | 36:0 |
|  | 15-24 weeks | - | - | - | - |  | - |

Supplement 5. Number of positive and negative PCR-confirmed SARS-CoV-2 test results in individuals tested for SARS-CoV-2 for 12-17-year-olds in England by vaccination status, interval after vaccination and previous infection status. Data is shown for the omicron variant.

|  |  | Previous infection (before vacc) | | | | |  | Previous infection (after vacc) | |
| --- | --- | --- | --- | --- | --- | --- | --- | --- | --- |
|  |  | None | Wild type | Alpha | Delta | Omicron |  | Delta | Omicron |
| Vaccination status | Time after vaccination | case:control | case:control | case:control | case:control | case:control |  | case:control | case:control |
| Unvaccinated |  | 53358:70999 | 2100:2039 | 3582:3143 | 12108:10511 | 109:106 |  | 12108:10511 | 109:106 |
| Dose 1 - Pfizer | 0-1 week | 3950:3160 | 160:51 | 234:62 | 1190:420 | 4:1 |  | 0:0 | 0:0 |
|  | 2-14 weeks | 63120:60277 | 2200:465 | 3567:912 | 12900:4807 | 12:6 |  | 68:56 | 3:4 |
|  | 15-24 weeks | 7485:10832 | 291:152 | 483:301 | 1007:691 | 0:0 |  | 483:330 | 82:27 |
|  | 25-39 weeks | 280:574 | 13:10 | 21:17 | 19:21 | 0:0 |  | 85:50 | 24:6 |
|  | 40 weeks | 17:28 | 1:1 | 1:2 | 0:0 | 0:0 |  | 3:3 | 0:0 |
| Dose 2 - Pfizer | 0-1 week | 8267:6871 | 297:71 | 414:115 | 1425:490 | 3:0 |  | 305:88 | 30:3 |
|  | 2-14 weeks | 28899:22987 | 939:340 | 1603:559 | 4142:1589 | 1:0 |  | 1633:526 | 33:7 |
|  | 15-24 weeks | 2161:2772 | 74:65 | 128:93 | 90:56 | 0:1 |  | 136:99 | 21:2 |
|  | 25-39 weeks | 1129:1498 | 40:44 | 64:51 | 2:3 | 4:5 |  | 89:47 | 2:1 |
|  | 40 weeks | 56:100 | 1:1 | 1:3 | 0:0 | 1:0 |  | 5:7 | 6:1 |
| Booster - Any mRNA vaccine | 0-1 week | 1000:1045 | 27:13 | 40:17 | 58:16 | 2:1 |  | 67:22 | 4:0 |
|  | 2-14 weeks | 2678:2384 | 80:41 | 104:52 | 71:36 | 7:3 |  | 216:53 | 9:2 |
|  | 15-24 weeks | 102:169 | 4:3 | 5:6 | 0:1 | 1:1 |  | 9:6 | 1:0 |

1.	UK Health Security Agency. SARS-CoV-2 variants of concern and variants under investigation in England. Technical briefing 44. 2022. <https://assets.publishing.service.gov.uk/government/uploads/system/uploads/attachment_data/file/1093275/covid-technical-briefing-44-22-july-2022.pdf> (accessed 17th August 2022).
